# Fusion of Imaging and Non-Imaging Data for Disease Trajectory Prediction for COVID-19 Patients

**DOI:** 10.1101/2021.12.02.21267211

**Authors:** Amara Tariq, Siyi Tang, Hifza Sakhi, Leo Anthony Celi, Janice M. Newsome, Daniel L. Rubin, Hari Trivedi, Judy Wawira Gichoya, Imon Banerjee

## Abstract

**Purpose:** This study investigates whether graph-based fusion of imaging data with non-imaging EHR data can improve the prediction of disease trajectory for COVID-19 patients, beyond the prediction performance of only imaging or non-imaging EHR data.

**Materials and Methods:** We present a novel graph-based framework for fine-grained clinical outcome prediction (discharge, ICU admission, or death) that fuses imaging and non-imaging information using a similarity-based graph structure. Node features are represented by image embedding and edges are encoded with clinical or demographic similarity.

**Results:** Our experiments on data collected from Emory Healthcare network indicate that our fusion modeling scheme performs consistently better than predictive models using only imaging or non-imaging features, with f1-scores of 0.73, 0.77, and 0.66 for discharge from hospital, mortality, and ICU admission, respectively. External validation was performed on data collected from Mayo Clinic. Our scheme highlights known biases in the model prediction such as bias against patients with alcohol abuse history and bias based on insurance status.

**Conclusion:** The study signifies the importance of fusion of multiple data modalities for accurate prediction of clinical trajectory. Proposed graph structure can model relationships between patients based on non-imaging EHR data and graph convolutional networks can fuse this relationship information with imaging data to effectively predict future disease trajectory more effectively than models employing only imaging or non-imaging data. Forecasting clinical events can enable intelligent resource allocation in hospitals. Our graph-based fusion modeling frameworks can be easily extended to other prediction tasks to efficiently combine imaging data with non-imaging clinical data.

## INTRODUCTION

To successfully perform any clinical prediction task, it is essential to learn effective representations of various data captured during patient encounters and model their interdependencies, including patient demographics, diagnostic codes, and radiologic imaging. Graph convolutional neural networks (GCN) present an intuitive and elegant way of processing multi-modal data presented as a graph structure. Recent works in GCN have enabled fusion of various data types and preserve their inter-dependency including imaging data such as MRI, CT scans, or X-rays with non-imaging electronic health record (EHR) or phenotypic data. Availability of comprehensive public databases such as TADPOLE [1], Alzheimer’s Disease Neuroimaging Initiative (ADNI)^1^ and ABIDE^2^ containing imaging and non-imaging information for patients with Alzheimer and autism spectrum disorder (ASD), respectively, has facilitated the use of GCN for disease prediction [2, 3, 4, 5, 6] with innovation in GCN architecture involving kernel size selection and use of recurrence. We build on this trend of research by applying the GCN model to understand the relationship between imaging and non-imaging data and by incorporating holistic weighted edge formation based on patient clinical history and demographic information. We propose a novel framework for fine-grained clinical event prediction for COVID-19 patients based on GCN-inspired models.

## BACKGROUND AND SIGNIFICANCE

Much of the literature regarding predictive modeling for COVID-19 patients has been largely focused on either diagnosis by processing imaging data (chest X-rays) through deep convolutional neural networks (CNN) [7-12], or mortality prediction using clinical risk factors [13-17]. In contrast, we propose a graph-based framework (see Figure 4) for predictive modeling of disease trajectory during hospitalization of the patient. Our framework includes multi-stage predictive models for various clinical events such as discharge from the hospital, ICU admission, and mortality. While most of the previous work has focused on either imaging data or hand-picked clinical factors [15, 16, 18-20], our framework fuses chest radiographs with non-imaging EHR data to capture patient similarity based on clinical history and demographic factors. While a pre-trained model was used for chest X-ray featurization, we developed innovative feature engineering schemes to model sparse information regarding past medical procedures and diagnosed illnesses of patients. Our relational graph-based modeling allows prediction processes to gather cues from similar cases, i.e., patients with similar demographic features and medical history. Our experiments clearly indicate merits of our fusion modeling in comparison to models employing only imaging or only non-imaging features. Biases based on demographic features like race, or socio-economic indicators like insurance status, or alcohol use, may affect clinical decision making regarding discharge from hospital or admission into the ICU [21, 22]. Therefore, we investigated disparity in the predictions made by the proposed framework.

## MATERIALS AND METHODS

We developed a graph-based fusion AI model to predict COVID-19 disease trajectory by using multi-modal patient data including radiology imaging data, demographic information, and clinical history (Figure 1).

**Figure 1:**
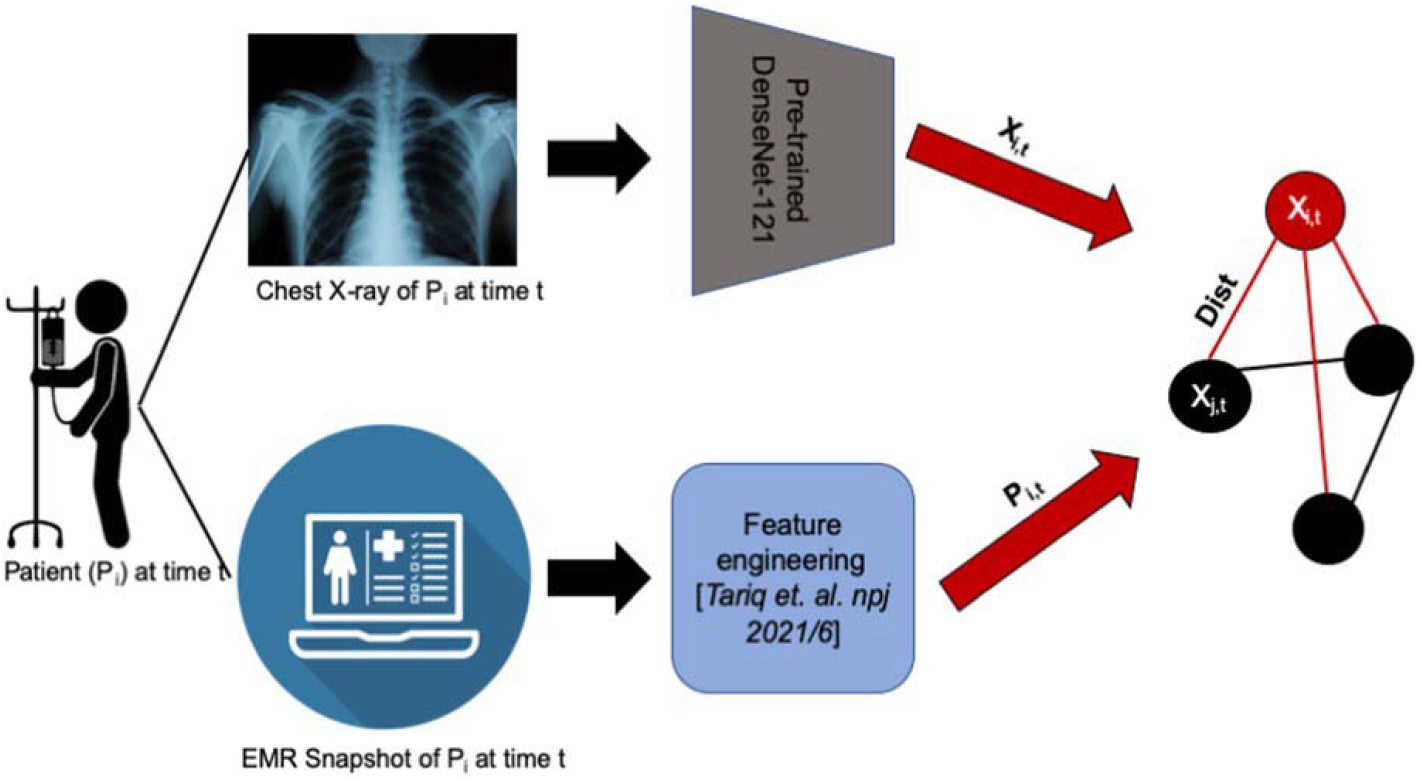
Proposed Graph-based fusion AI framework for modeling image and EHR data

### Cohort description

Following approval of the Emory Institutional Review Board (IRB), we collected all chest X-rays of patients with at least one positive RT-PCR test for COVID-19, performed in 12 centers of the Emory Healthcare network from January 2020 to December 2020. As shown in Figure 2, the data consisted of 47,555 chest X-rays belonging to 23,831 unique patients. We only considered posteroanterior (PA) and anteroposterior (A) view of chest radiographs during the period of hospitalization. We also collected hospital admission, mortality and discharge data from the hospital billing system. Patients who were never admitted, or those who were admitted but did not receive regular intervals of x-rays (at minimum every three days) were discarded. This left 2,201 unique patients corresponding to 2,983 hospitalizations for which chest-x rays were obtained at least every 3 days (7,813 total chest-x rays). In order to avoid data leakage, we applied a patient-wise split of the train and test sets. Some patients were hospitalized multiple times during the year 2020. We considered the chest X-ray exam date and time as a timepoint for predicting future clinical events and stratified these events based on severity – discharged from hospital with 3 days, admission to ICU, or death. There were 929 patients and 4,850 chest X-rays after which the patient was admitted to the hospital for more than three days. Out of these, 380 patients corresponding to 543 chest Xrays were admitted to ICU within 3 days. There were 1,754 patients corresponding to 2,963 chest X-rays who were discharged from hospital within 3 days. 220 patients corresponding to 502 chest X-rays died within 3 days. The selection process for this cohort is shown in Figure 2. Outcome data for this cohort was collected from hospital billing records and warehouse EHR data sources. Our goal was to predict adverse events as early as possible to provide enough decision making time for the clinicians. In our preliminary experiments, we evaluated 3 days up to 7 days, however given the unbalanced dataset, we found 3 days to be the most prevalent in terms of different prediction labels. This cut-off provided a rather balanced distribution of discharged vs. not discharged labels.

**Figure 2:**
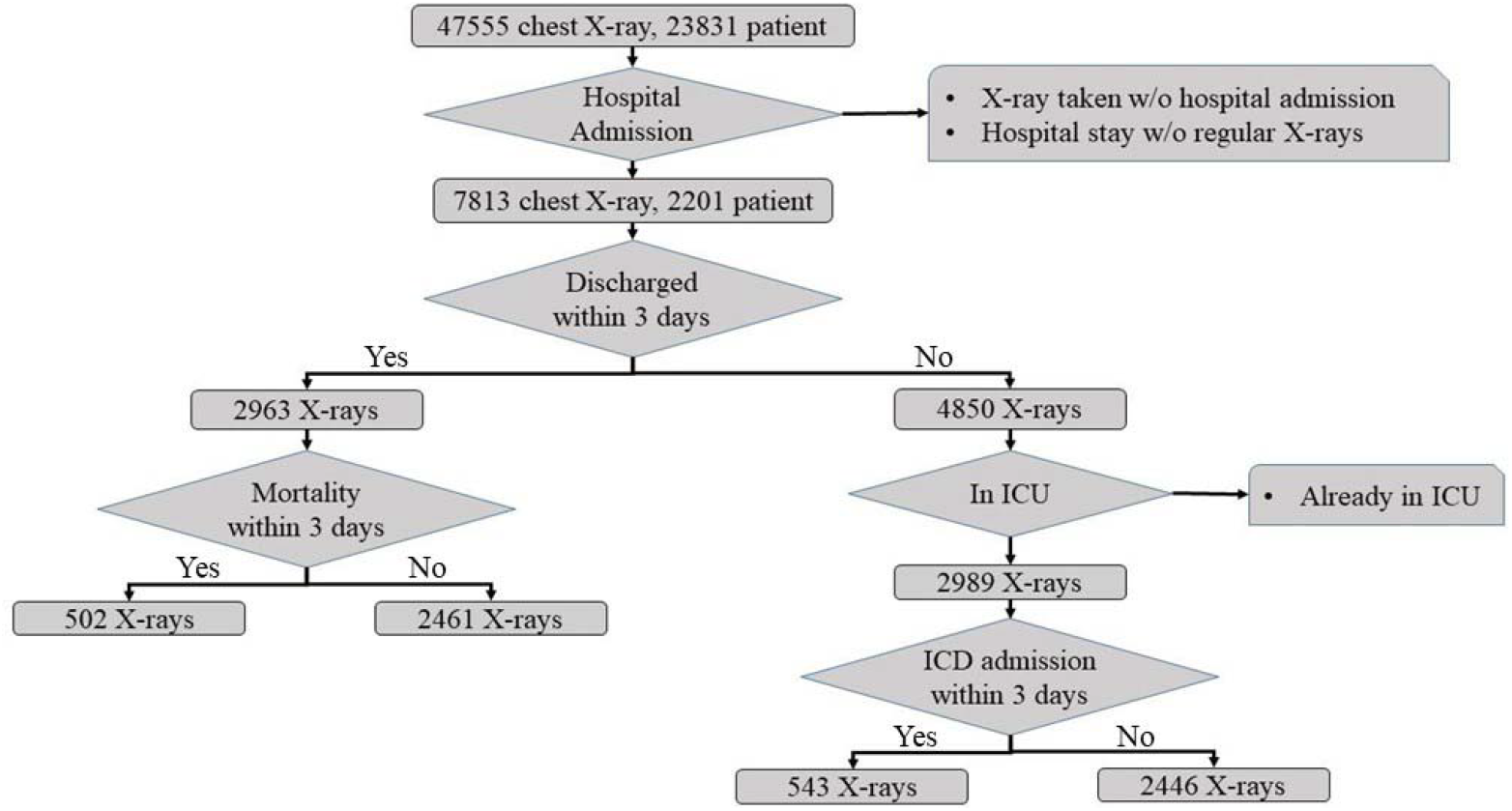
CONSORT style diagram for Cohort selection for internal data

### External test

With the approval of Mayo Institutional Review Board, we shipped our Emory trained model to Mayo and evaluated externally on 50 unique patients admitted to the Mayo Clinic hospital between Jan 2020 – Dec, 2020 with a positive RT-PCR test. The patients have 293 chest X-rays during the period of hospitalization. Demographic and comorbidities statistics of both cohorts are provided in Table 1.

**Table 1:** Cohort characteristics - demographics, clinical history and insurance status. The numbers have been represented in terms of complete dataset (total cohort) as well as train and test split.

| Characterization |  | Total cohort (2201) | Train (1762) | Test (439) | External test (50) |
| --- | --- | --- | --- | --- | --- |
| Gender | Male | 1111 | 887 | 224 | 34 |
|  | *Mean Age (std. deviation)* | *59.7(16.2)* | *59.2 (16.1)* | *61.5(16.3)* | *64.5 (16.3)* |
|  | Female | 1090 | 875 | 215 | 16 |
|  | *Mean Age (std. deviation)* | *59.0 (18.8)* | *58.7(18.9)* | *60.2(18.4)* | *59.8 (15.9)* |
| Race | African American | 1405(63.8%) | 1135(64.4%) | 270(61.5%) | 6 (12%) |
|  | Caucasian | 554(25.2%) | 427(24.2%) | 127(28.9%) | 43 (86%) |
|  | Asian | 51(2.3%) | 42(2.4%) | 9(2.1%) | 1(2%) |
|  | American Indian or Alaskan | 11(0.5%) | 7(0.4%) | 4(0.9%) | -- |
|  | Multiple | 6(0.3%) | 6(0.3%) | 0(0%) | -- |
|  | Native Hawaiian or Other | 6 (0.3%) | 4(0.2%) | 2(0.4%) | -- |
|  | Unknown | 168(7.6%) | 141(8.0%) | 27(6.2%) | -- |
| Ethnicity | Hispanic | 163(7.4%) | 133(7.5%) | 30 (6.8%) | -- |
|  | Non-Hispanic | 1894(86.1%) | 1517(86.1%) | 377(85.9%) | 50 (100%) |
|  | Unknown | 144(6.5%) | 112(6.4%) | 32(7.3%) | -- |
| Comorbidities | Diabetes | 1187(53.9%) | 937 (53.2%) | 250(56.9%) | 7(14%) |
|  | Renal Disease | 1330(60.4%) | 1050(59.6%) | 280(63.8%) | 10(20%) |
|  | Hypertension | 1711(77.7%) | 1356(77.0%) | 355(80.9%) | 19 (38%) |
|  | Respiratory Disease | 1948(88.5%) | 1556(88.3%) | 392(89.3%) | 27 (54%) |
| Insurance Status | Medicare | 1034(47.0%) | 806(45.7%) | 228(51.9%) | Not available |
|  | Commercial | 784(35.6%) | 651 (36.9%) | 133(30.1%) |  |
|  | Medicaid | 125(5.7%) | 100(5.7%) | 25(5.7%) |  |
|  | Others | 258 (11.7%) | 205(11.6%) | 53(12.1%) |  |
| Alcohol Use | No | 1464(66.5%) | 1178(66.9%) | 286(65.2%) |  |
|  | Yes | 245(11.1%) | 213(12.1%) | 32(7.3%) |  |
|  | Not Reported | 492(22.4%) | 371 (21.1%) | 121(27.6%) |  |

### Multimodal encounter data description and feature engineering

To fulfill the aim of multi-modal data fusion, we tested different approaches to handling imaging and non-imaging data.

#### Imaging Data

We used a DenseNet-121 [23] pre-trained on the open-source chest CheXpert X-ray dataset [24], and fine-tuned on 199,029 non-COVID chest X-rays from EUH acquired in 2019, for processing normalized images. We dropped the final softmax classification layer of the DenseNet-121 model and extracted 1024-dimensional feature vectors from each image to construct a dense representation of the images.

#### Non-Imaging EHR Data

We extracted the current and historic EHR data from 2020 for all patients in our cohort.

#### Demographic information

Gender (male/female), self-reported race (African American, Caucasian, Native Hawaiian or Other Pacific Islander, Asian, American Indian or Alaska Native, Multiple, Unknown), ethnic group (Hispanic or Latino, Non-Hispanic or Latino, Unknown), and age at the time of admission (binned in 10-year intervals). All features were one-hot encoded.

#### Current Procedural Terminology codes (CPT)

CPT is a five-digit procedure code that reports medical, surgical, and diagnostic procedures and services to entities such as physicians, health insurance companies and accreditation organizations. CPT codes are maintained and grouped in a hierarchical structure by the American Medical Association (AMA). Each CPT code was reduced into a higher-order parent category based on the defined hierarchy ^3^ (details available in supplementary material). We selected all of 21 groups with more than 1000 occurrences.

#### *Comorbiditie*s

Past and current diagnoses of patients are structured as International Classification of Disease, 9th edition (ICD-9) codes which we grouped based on hierarchical structure [22] (details available in supplementary material). We selected all of 29 groups occurring more than 5000 times in the data.

### Graph-based Fusion model - Image and EHR

Like any CNN, a GCN learns to generate a vector embedding of each node optimized for the downstream prediction task, While CNN defines ‘neighborhood’ of an instance based on spatial proximity such as neighboring pixels in images, GCN allows the neighborhood to be defined by edges in graph structure. Edges can be defined by any relevant similarity metrics/interconnection between the nodes, providing extreme flexibility for modeling complex clinical scenarios. Node embeddings generated by a GCN are dependent on node representation itself and neighborhood/edge-connected nodes. In our graph design, chest X-ray embeddings (1024-dimensional vector) are used as nodes and edges are decided by similarity in patients’ demographic information or medical history (see Figure 3). Therefore, our GCN supports the notion of ‘neighborhoods’ based on clinical and/or demographic similarity between patients and makes predictions for future clinical events by considering trends among similar patients.

**Figure 3:**
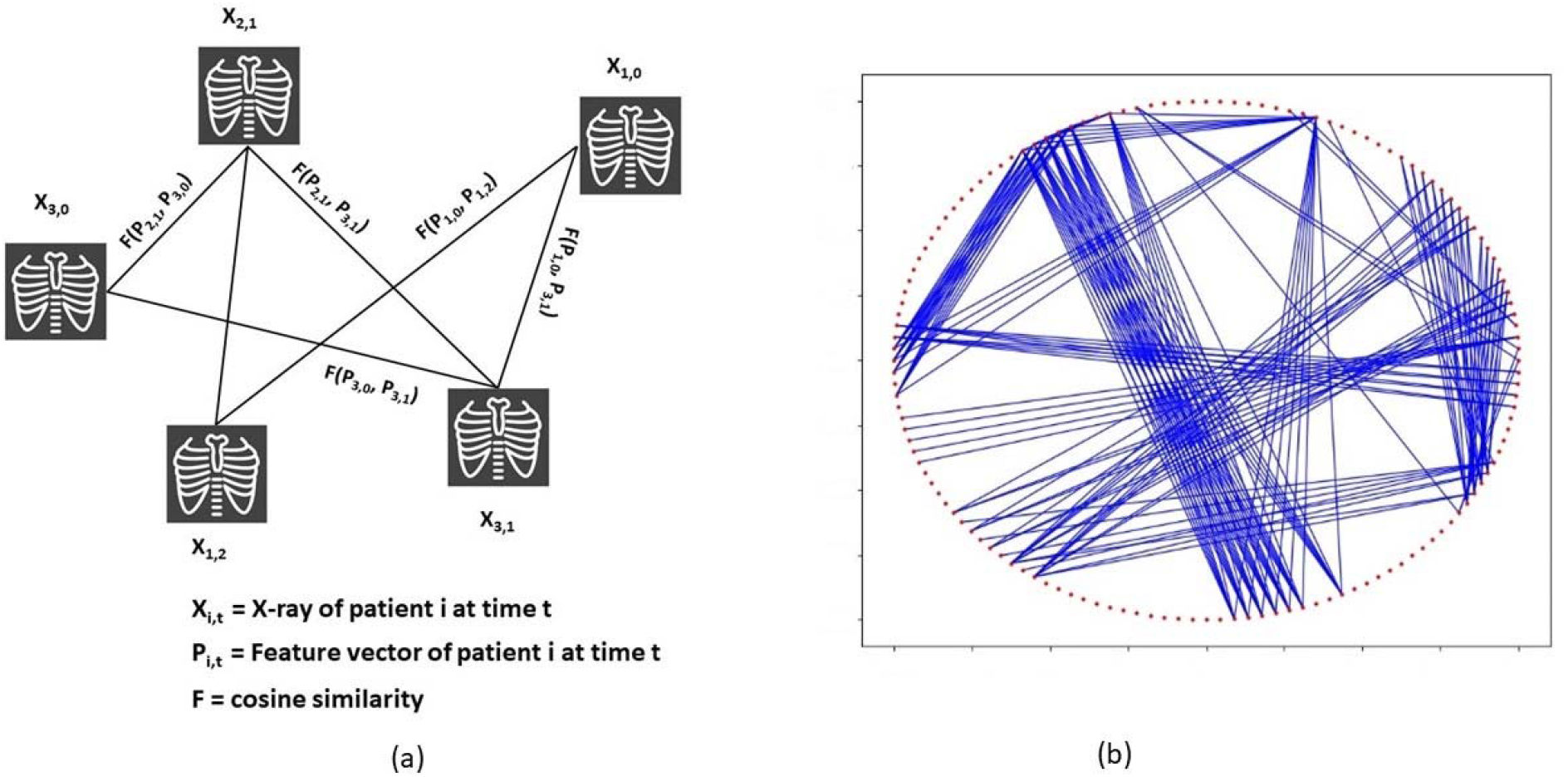
Graph formation based on chest X-rays and patient similarity information; a) sample graph construction, b) 1% random subsample of an actual graph – red dots are nodes indicated by chest X-rays in sample graph, blue lines are edges which are constructed between nodes when similarity between corresponding patient features exceed a certain threshold.

**Figure 4:**
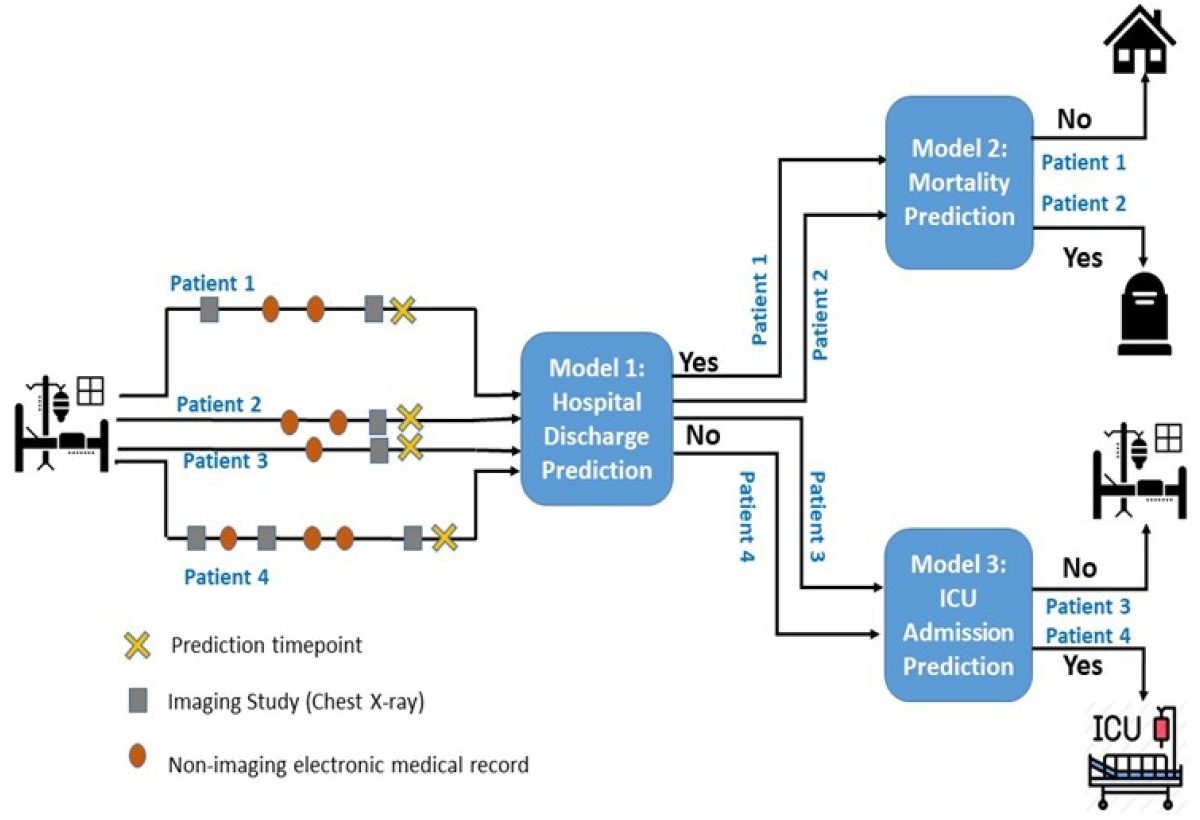
Branched framework for disease trajectory prediction with three sequential decision points (highlighted in blue box)

For edge formation, we encoded an EHR feature vector for each node (chest radiograph) based on one-hot encoding of EHR information of the patient corresponding to that radiograph. Edge formulation only employs retrospective data collected before the chest radiograph exam date to avoid any data leakage. Cosine similarity between EHR feature vectors corresponding to two nodes is used to decide edge between the nodes, while edge weight encodes the strength of the similarity between nodes.

Many GCN variations are designed for transductive learning such that they can only process graph structures used for training with no ability to generalize to unseen nodes or new graph structure. To avoid this limitation, we used the SAGE (SAmple and aggreGatE) graph convolution network (GraphSAGE) [25] that optimizes a sample aggregate function to collect ‘messages’ from neighboring nodes while generating vector embedding of a node. For inference, GraphSAGE employs an optimized aggregate function to generate embedding for unseen nodes in unseen graph structures. GraphSAGE based prediction models can inductively reason to assign predictive labels to unlabeled nodes by learning from labelled nodes in the graph.

#### Branched Framework of Prediction

Forecasting the trajectory of disease, in terms of three clinical events, once the patient has been hospitalized is the focus of this work. Given the challenge of collecting a balanced dataset for multi-class classification, we modeled 3 sequential decision points and developed a pipeline for comprehensive prediction of possible clinical events. (Model1) - Prediction of discharge from the hospital: In the first decision point, Model-1 predicts whether the patient will stay in the hospital for more than 3 days (positive label) or not. (Model-2) - Mortality prediction: For negatively labeled instances by Model-1, in the second decision point, Model-2 predicts whether the patient will expire within 3 days (positive label) or not. (Model-3) - Prediction of Admission in ICU: Finally, for positively labelled instances from Model-1, Model-3 predicts whether the patient will be admitted to ICU within 3 days (positive label) or not. Distribution of positive and negative class labels is highly imbalanced, especially for Model-2 and Model-3 (Figure 2). We employed undersampling of majority label and weighted loss to tackle this challenge.

In this framework, disease trajectory is predicted as predictive events include indicators of worsening (admission in ICU, mortality) or improving (discharge from hospital) prognosis. A patient is evaluated every time a chest radiograph is taken, while staying in the hospital.

#### Model comparison and statistical evaluation

Evaluation of the proposed modeling framework is focused on two aspects; 1) performance of the fusion model in comparison to models using single modality (either imaging data or non-imaging data), 2) comparative effectiveness of different EHR data sources in terms of graph structure definition-based prediction. We designed a Random Forest classifier and used the Pulmonary X-ray severity (PXS) scores computed by a CNN [26] to predict clinical events as well. As PXS scores are based on deep learning based processing of imaging data, we wanted to establish the benefits of our selection of imaging features in our relational graph by comparison. We report the model performance in terms of Area Under the Receiver Operating Characteristics curve (AUCROC), precision, recall and f1-score on held-out test-set from Emory University cohort, as well as on external test-set composed of patients’ of Mayo Clinic.

## RESULTS

### Quantitative performance

Our framework consists of three binary prediction models: Model-1 (Discharge from hospital), Model-2 (mortality), Model-3 (ICU admission). Tables 2 presents results for these three prediction models evaluated using the class-wise and aggregated (weighted average) precision, recall, and F-score as well as confidence interval (95% confidence) on a randomly held-out set of test samples. In Figure 5, we also represent the receiver operating characteristics (ROC) curves for these evaluations.

**Table 2:**
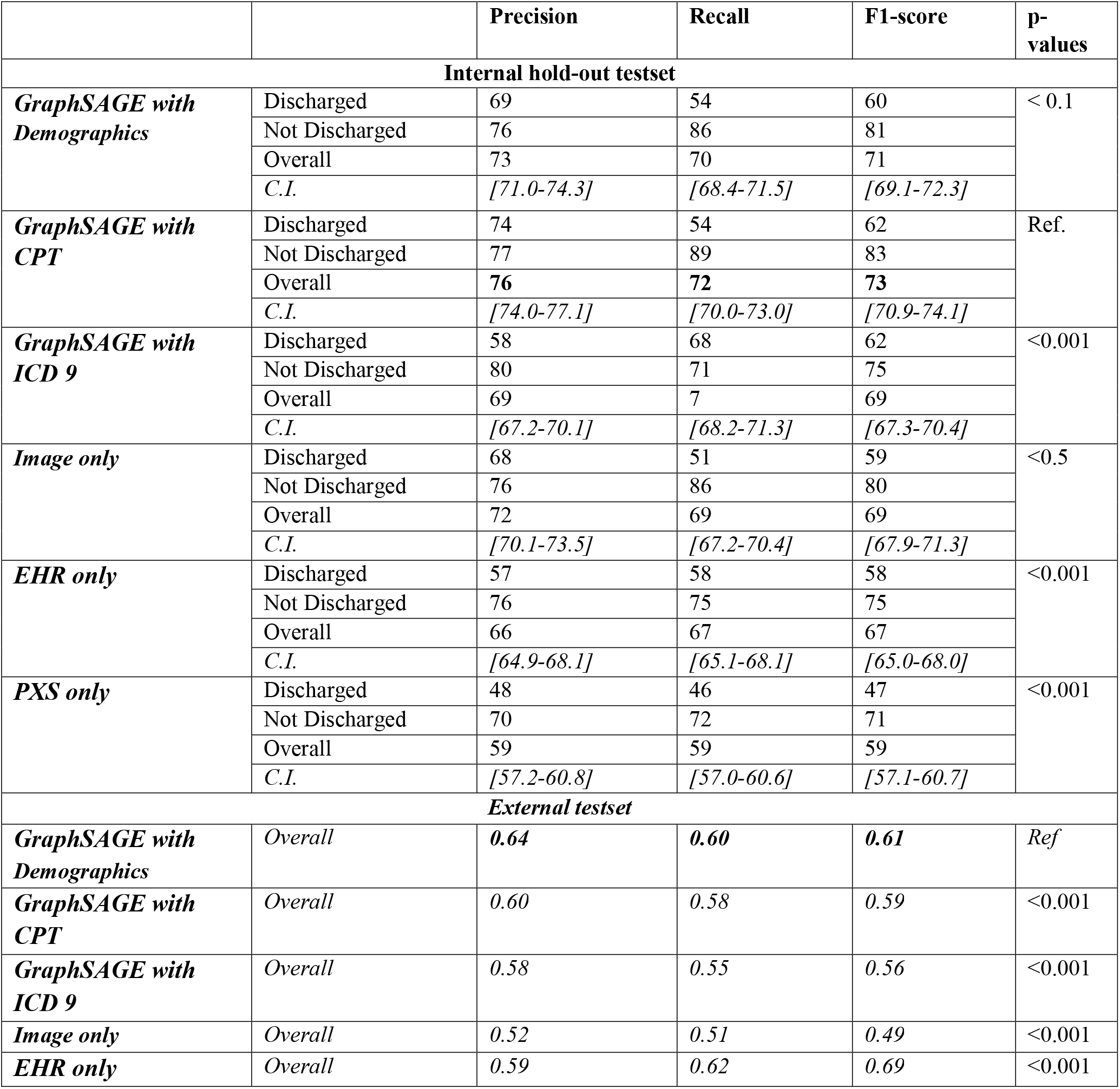
Performance of Model 1. Prediction of discharge from hospital within 3 days. Total number of held-out test samples - 1,545 with 563 discharged patients. CI stands for 95% confidence range calculated using bootstrapping with 1000 randomly subsampled sets from the held-out test set. Optimal performance is highlighted in bold. p-values have been computed using pairwise t-test with GraphSAGE CPT. Overall performance of the external dataset was represented in terms of macro average of precision, recall, and F-score.

|  |  | Precision | Recall | F1-score | p-values |
| --- | --- | --- | --- | --- | --- |
| Internal hold-out testset |  |  |  |  |  |
| GraphSAGE with Demographics | Discharged | 69 | 54 | 60 | < 0.1 |
|  | Not Discharged | 76 | 86 | 81 |  |
|  | Overall | 73 | 70 | 71 |  |
|  | C.I. | [71.0-74.3] | [68.4-71.5] | [69.1-72.3] |  |
| GraphSAGE with CPT | Discharged | 74 | 54 | 62 | Ref. |
|  | Not Discharged | 77 | 89 | 83 |  |
|  | Overall | 76 | 72 | 73 |  |
|  | C.I. | [74.0-77.1] | [70.0-73.0] | [70.9-74.1] |  |
| GraphSAGE with ICD 9 | Discharged | 58 | 68 | 62 | <0.001 |
|  | Not Discharged | 80 | 71 | 75 |  |
|  | Overall | 69 | 7 | 69 |  |
|  | C.I. | [67.2-70.1] | [68.2-71.3] | [67.3-70.4] |  |
| Image only | Discharged | 68 | 51 | 59 | <0.5 |
|  | Not Discharged | 76 | 86 | 80 |  |
|  | Overall | 72 | 69 | 69 |  |
|  | C.I. | [70.1-73.5] | [67.2-70.4] | [67.9-71.3] |  |
| EHR only | Discharged | 57 | 58 | 58 | <0.001 |
|  | Not Discharged | 76 | 75 | 75 |  |
|  | Overall | 66 | 67 | 67 |  |
|  | C.I. | [64.9-68.1] | [65.1-68.1] | [65.0-68.0] |  |
| PXS only | Discharged | 48 | 46 | 47 | <0.001 |
|  | Not Discharged | 70 | 72 | 71 |  |
|  | Overall | 59 | 59 | 59 |  |
|  | C.I. | [57.2-60.8] | [57.0-60.6] | [57.1-60.7] |  |
| External testset |  |  |  |  |  |
| GraphSAGE with Demographics | Overall | 0.64 | 0.60 | 0.61 | Ref |
| GraphSAGE with CPT | Overall | 0.60 | 0.58 | 0.59 | <0.001 |
| GraphSAGE with ICD 9 | Overall | 0.58 | 0.55 | 0.56 | <0.001 |
| Image only | Overall | 0.52 | 0.51 | 0.49 | <0.001 |
| EHR only | Overall | 0.59 | 0.62 | 0.69 | <0.001 |

**Figure 5:**
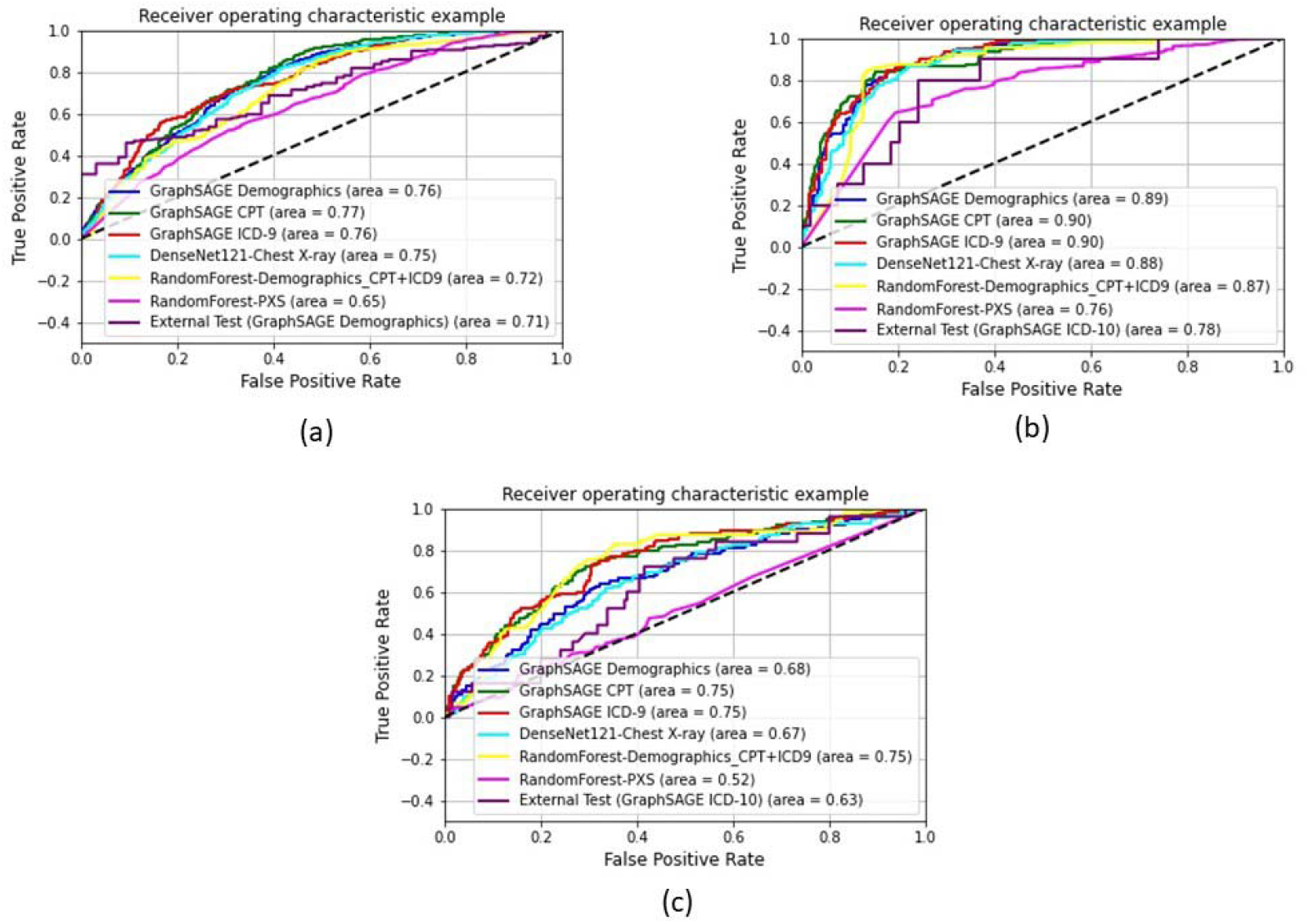
ROC curves; (a) Model-1: discharge prediction, (b) Model-2: Mortality Prediction, (c) Model-3: ICU Admission Prediction

Tables 2, 3, and 4 show the performance of predictive models - (1) *EHR only:* Random forest model using all EHR data sources as input (demographics, CPT groups, ICD-9 groups), (2) *PXS only:* Random forest model using only PXS score derived from the images as input, (3) *Image only*: softmax classification of the pretrained DensenNET-121 using chest X-ray as input, and (4) *GraphSAGE with XX:* GraphSAGE network with graph structure based on different EHR sources (XX) like demographics, CPT groups, and ICD-9 groups.

**Table 3:** Performance Model 2. Prediction of mortality of patients within 3 days. Total number of held-out test samples - 563 with 445 alive patients. CI stands for 95% confidence range calculated using bootstrapping with 1000 randomly subsampled sets from the held-out test set. Optimal performance is highlighted in bold. p-values have been computed using pairwise t-test with GraphSAGE CPT. Overall performance of the external dataset was represented in terms of macro average of precision, recall, and F-score.

|  |  | Precision | Recall | F1-score | p-values |
| --- | --- | --- | --- | --- | --- |
| Internal hold-out testset |  |  |  |  |  |
| GraphSAGE with Demographics | Alive | 87 | 96 | 91 | <0.001 |
|  | Dead | 73 | 45 | 55 |  |
|  | Overall | 80 | 70 | 73 |  |
|  | C.I. | [76.4-83.2] | [67.2-73.3] | [70.0-76.4] |  |
| GraphSAGE with CPT | Alive | 91 | 92 | 91 | Ref. |
|  | Dead | 68 | 65 | 67 |  |
|  | Overall | 80 | 79 | 79 |  |
|  | C.I. | [76.6-82.6] | [75.6-81.7] | [76.2-81.8] |  |
| GraphSAGE with ICD 9 | Alive | 91 | 90 | 90 | >0.5 |
|  | Dead | 63 | 64 | 64 |  |
|  | Overall | 77 | 77 | 77 |  |
|  | C.I. | [73.9-79.9] | [74.3-80.7] | [74.1-79.7] |  |
| Image only | Alive | 90 | 89 | 89 | <0.5 |
|  | Dead | 60 | 63 | 61 |  |
|  | Overall | 75 | 76 | 75 |  |
|  | C.I. | [72.3-78.2] | [72.7-79.2] | [72.5-78.4] |  |
| EHR only | Alive | 78 | 51 | 62 | <0.001 |
|  | Dead | 88 | 96 | 92 |  |
|  | Overall | 83 | 74 | 77 |  |
|  | C.I. | [79.8-86.3] | [70.5-76.6] | [73.6-80.0] |  |
| PXS only | Alive | 85 | 87 | 86 | <0.001 |
|  | Dead | 47 | 42 | 44 |  |
|  | Overall | 66 | 64 | 65 |  |
|  | C.I. | [62.2-69.0] | [61.3-67.7] | [61.7-68.1] |  |
| External testset |  |  |  |  |  |
| GraphSAGE with Demographics | Overall | 0.55 | 0.59 | 0.54 | <0.1 |
| GraphSAGE with CPT | Overall | 0.77 | 0.59 | 0.61 | <0.1 |
| GraphSAGE with ICD 9 | Overall | 0.63 | 0.64 | 0.63 | Ref |
| Image only | Overall | 0.62 | 0.74 | 0.57 | >0.5 |
| EHR only | Overall | 0.81 | 0.63 | 0.66 | <0.001 |

**Table 4:** Performance Model 3. Prediction of admission to the ICU. Total number of held-out test samples - 592 with 110 ICU admitted patients.CI stands for 95% confidence range calculated using bootstrapping with 1000 randomly subsampled sets from the held-out test set. p-values have been computed using pairwise t-test with GraphSAGE ICD. Overall performance of the external dataset was represented in terms of macro average of precision, recall, and F-score.

|  |  | Precision | Recall | F1-score | p-values |
| --- | --- | --- | --- | --- | --- |
| **GraphSAGE with Demographics** | Not Admitted to ICU | 87 | 74 | 80 | <0.001 |
|  | Admitted to ICU | 32 | 53 | 39 |  |
|  | Overall | 59 | 63 | 60 |  |
|  | C.I. | [56.8-62.2] | [59.8-66.9] | [56.7-62.9] |  |
| **GraphSAGE with CPT** | Not Admitted to ICU | 89 | 79 | 83 | <0.01 |
|  | Admitted to ICU | 38 | 57 | 45 |  |
|  | Overall | 63 | 68 | 64 |  |
|  | C.I. | [60.9-66.0] | [64.8-71.4] | [61.7-67.4] |  |
| **GraphSAGE with ICD 9** | Not Admitted to ICU | 88 | 85 | 86 | Ref. |
|  | Admitted to ICU | 43 | 50 | 46 |  |
|  | Overall | **65** | **67** | **66** |  |
|  | C.I. | [62.0-68.4] | [63.8-70.8] | [62.7-69.2] |  |
| **Image only** | Not Admitted to ICU | 86 | 75 | 80 | <0.01 |
|  | Admitted to ICU | 30 | 48 | 37 |  |
|  | Overall | 58 | 61 | 59 |  |
|  | C.I. | [55.9-60.9] | [58.3-64.9] | [55.8-61.6] |  |
| **EHR only** | Not Admitted to ICU | 84 | 91 | 87 | <0.001 |
|  | Admitted to ICU | 38 | 25 | 30 |  |
|  | Overall | 61 | 58 | 59 |  |
|  | C.I. | [57.1-65.1] | [55.0-60.6] | [55.5-61.7] |  |
| **PXS only** | Not Admitted to ICU | 82 | 81 | 82 | <0.01 |
|  | Admitted to ICU | 21 | 22 | 21 |  |
|  | Overall | 52 | 52 | 52 |  |
|  | C.I. | [48.6-54.4] | [48.6-54.4] | [48.6-54.4] |  |
| **External testset** |  |  |  |  |  |
| **GraphSAGE with Demographics** | Overall | 0.51 | 0.51 | 0.51 | <0.5 |
| **GraphSAGE with CPT** | Overall | 0.54 | 0.56 | 0.54 | <0.001 |
| **GraphSAGE with ICD 9** | Overall | **0.56** | **0.54** | **0.55** | Ref |
| **Image only** | Overall | 0.52 | 0.53 | 0.51 | <0.001 |
| **EHR only** | Overall | 0.41 | 0.36 | 0.20 | <0.01 |

For both prediction of discharge from hospital and mortality, GraphSAGE with CPT achieved the highest performance, surpassing baseline single modality models - Image-only and EHR only, while PXS-only model also achieved suboptimal performance. GraphSAGE with demographics and ICD do not provide any significant boost over the baseline performance for both the prediction tasks. However, GraphSAGE with ICD was the best performing model for ICU admission prediction, surpassing the performance of single modality models and PXS-only model.

Overall results demonstrate that graph-based models increase the performance beyond the performance of any individual source for all the target prediction tasks. The performance tables show p-values, computed by statistical t-test to evaluate statistically significant performance differences, for all models at all decision points compared with the best performing model at the corresponding decision point.

### Model bias analysis

Interestingly, GraphSAGE models trained on demographic similarity data, do not surpass the performance of the baseline models, even for the ICU admission and mortality. We used the Aequitas toolkit [27] to analyze model disparity focused on false positive rate (FPR) and false omission rates (FOR). Figure 6 shows these FPR and FOR disparity plots based on alcohol use, race and insurance status for discharge (Model 1) and ICU admission (Model 3). FPR represents the rate of false positive prediction when a certain subgroup is falsely predicted to have positive label where positive label is punitive in nature such as discharge from hospital. Similarly, FOR represents the fraction of cases from a certain subgroup who are falsely omitted from a positive prediction group where the positive label has an assistive nature such as admission to ICU. Disparity plots represent comparisons of FPR and FOR of different subgroups in reference to a reference subgroup. Adhering to convention, we chose the majority subgroup as the reference for both plots. Higher disparity values indicate unfavorable bias while lower values indicate favorable bias. Our analysis indicates that the model for discharge prediction is unfavorably biased towards patients with alcohol abuse history, Caucasian population. While Medicaid insurance has a negative bias towards discharge, commercial insurance has a high False Omission rate for ICU admission. No racial bias was identified in our predictive models. This conclusion is based on an acceptable bias range of 0.8 to 1.2.

**Figure 6:**
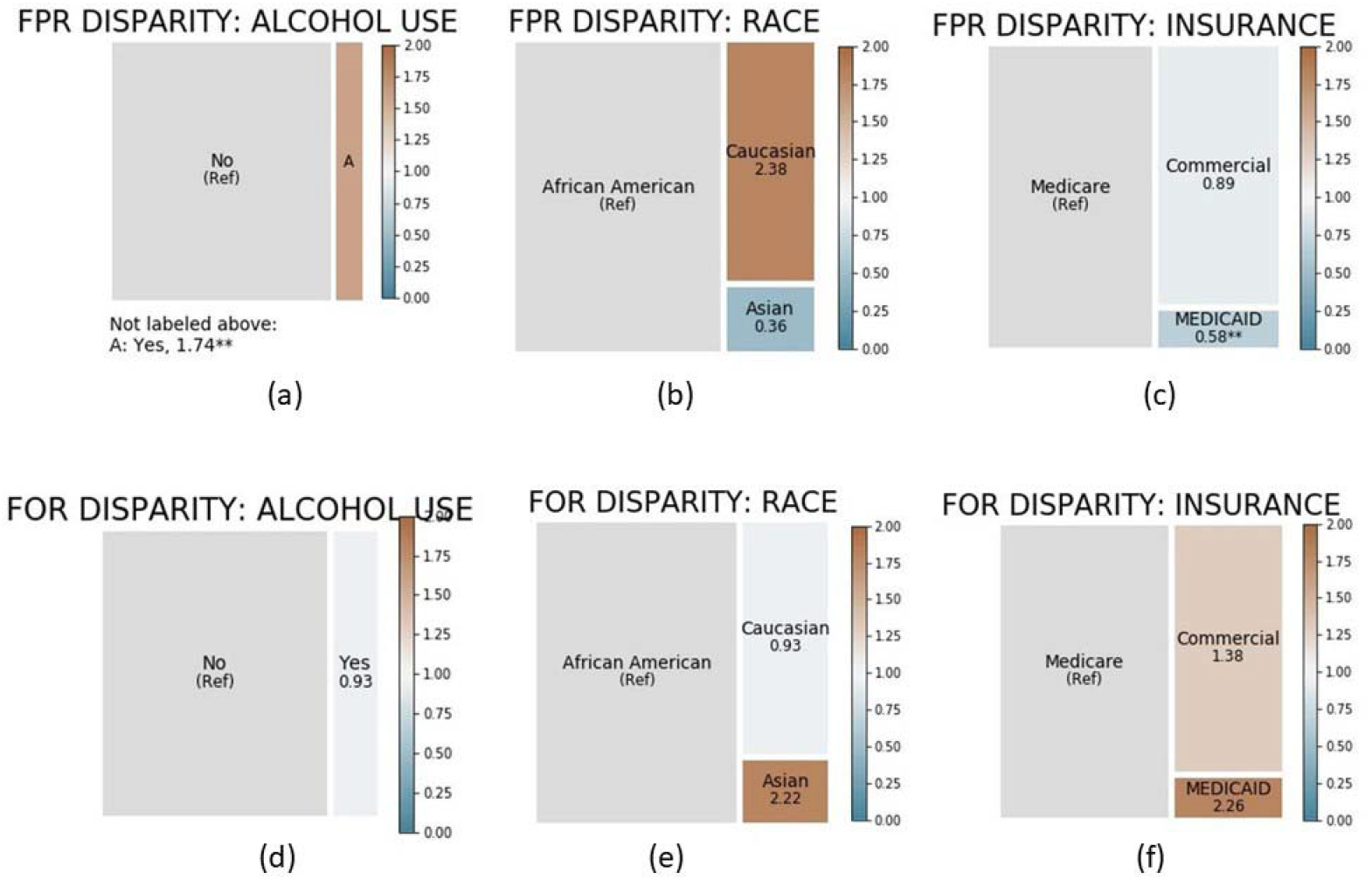
False Positive Rate (FPR) based disparity in Model-1: Discharge Prediction; (a) alcohol use based disparity, (b) race based disparity (c) insurance status based disparity. False Omission Rate (FOR) based disparity in Model-3: ICU Admission Prediction; (d) alcohol use based disparity, (e) race based disparity (f) insurance status based disparity

### External Validation

During the external validation, we found that procedures (CPT codes) and comorbidities (ICD-10) for the external population are very different from common codes of our internal validation dataset (lists of common CPT and ICD-10 codes are provided in supplementary material). Following the proposed strategy, we built graphs based on demographics (age, gender, race, and ethnicity), common ICD-10 and common CPT codes for this cohort and tested our GraphSAGE models as well as image-only and EHR-only models for all prediction labels. Results of their performance are provided in Table 2 - 4. It is clear that graph based models have higher and more consistent generalization capabilities than baseline models (image-only and EHR-only) achieving better F-score for all three tasks. Baseline models do achieve higher AUROC only for mortality prediction

## DISCUSSION

It appears that elements in clinical history, like past medical procedures and comorbidities play a larger role in clinical event prediction than demographic features like age, gender, and race. Though past medical procedures often produced better performance than coded comorbidities, past procedures are already correlated with specific comorbidities. In addition to single modality model, we also compared the different GraphSAGE models against a model trained on PXS score[26] where the PXS were directly computed from the X-ray images and the CNN model was pretrained on ∼160,000 images from CheXpert and transfer learning on 314 CXRs from patients with COVID-19. GraphSAGE model also outperformed the PXS only model with a statistically significant margin. We also validated the performance of GraphSAGE against the individual modality on an external institution where the common comorbidities (diabetics, renal disease) are rare (<20%) compared to our internal training data (>55%) and as a consequence the respective procedures are less frequently performed. In such distribution shift, often the machine learning model failed to generalize [28]; however graph based modeling is flexible enough to handle data-shift by updating the *similarity definition* for edge generation and produce comparable results on the external data.

Despite the encouraging performance, the bias analysis shows that model performance is biased towards race and current insurance status for discharge and ICU admission. However, this is more related towards the practice since as multiple past studies have highlighted biases in crucial care services and discharge delays based on insurance status [29, 30, 31, 32, 33, 34].

### Limitation of the study

The proposed framework may face limitations in terms of application scope as it requires imaging data to be collected on regular interval and was trained on data collected from highly integrated academic healthcare system. Prediction interval is also limited to 3 days which is still longer than most studies done in the past [18, 19]. Due to the two-fold informational fusion of the GraphSAGE which involves mathematically irreversible calculations, it is not feasible to apply traditional model interpretation techniques, and hard to explain the decision reason for the prediction of the each node classification.

## CONCLUSION

We proposed a graph-based framework to preserve interdependencies in multi-modal data, i.e., EHR and radiologic images, to predict future clinical events (e.g. discharge, ICU admission, and mortality) for the in-patients population tested positive for COVID-19 within 3 days of admission. During the graph-based learning, in theory, two-fold information fusion of node features and graph structure ensure that relevant features (nodes and graph structure) for the targeted task are amplified while similar non-relevant attributes are suppressed. To our knowledge, this is the first attempt to encode and learn the patient-wise similarity within imaging data using a GCN model and apply that for predictive modeling hospital resource optimization. Our experiments clearly establish superiority of relational graph based fusion modeling, over single-modality predictive models, in terms of prediction performance and adaptation to different populations.

## Data Availability

All data used in the study is confidential, and is not publicly available.

## ACKNOWLEDGMENT

We acknowledge Emory Healthcare Network and Mayo Clinic for providing the data for internal and external validation.

## Footnotes

1 adni.loni.usc.edu

2 https://fcon_1000.projects.nitrc.org/indi/abide/

3 https://www.aapc.com/codes/cpt-codes-range/

