## Supplementary Material for "Fusion of Imaging and Non-Imaging Data for Disease Trajectory Prediction for COVID-19 Patients"

### Feature Engineering:

#### CPT Codes:

CPT ontology is made up of groups of related CPT codes which comprise of subgroups of CPT. We used this grouping structure to map individual CPT codes to 212 CPT categories, made up of subgroups of CPT ontology. Each subgroup is defined by numerically minimum and maximum CPT codes as well as any alphabetic modifier used with the numerical code. Supplementary Table 1 shows all CPT groups used as features.

| Group | Subgroup | Low | High | Modifier |
| --- | --- | --- | --- | --- |
| Anesthesia | Anesthesia for Procedures on the Head | 100 | 222 |  |
|  | Anesthesia for Procedures on the Neck | 300 | 352 |  |
|  | Anesthesia for Procedures on the Thorax (Chest Wall and Shoulder Girdle) | 400 | 474 |  |
|  | Anesthesia for Intrathoracic Procedures | 500 | 580 |  |
|  | Anesthesia for Procedures on the Spine and Spinal Cord | 600 | 670 |  |
|  | Anesthesia for Procedures on the Upper Abdomen | 700 | 797 |  |
|  | Anesthesia for Procedures on the Lower Abdomen | 800 | 882 |  |
|  | Anesthesia for Procedures on the Perineum | 902 | 952 |  |
|  | Anesthesia for Procedures on the Pelvis (Except Hip) | 1112 | 1173 |  |
|  | Anesthesia for Procedures on the Upper Leg (Except Knee) | 1200 | 1274 |  |
|  | Anesthesia for Procedures on the Knee and Popliteal Area | 1320 | 1444 |  |
|  | Anesthesia for Procedures on the Lower Leg (Below Knee) | 1462 | 1522 |  |
|  | Anesthesia for Procedures on the Shoulder and Axilla | 1610 | 1680 |  |
|  | Anesthesia for Procedures on the Upper Arm and Elbow | 1710 | 1782 |  |
|  | Anesthesia for Procedures on the Forearm, Wrist, and Hand | 1810 | 1860 |  |
|  | Anesthesia for Radiological Procedures | 1916 | 1936 |  |
|  | Anesthesia for Burn Excisions or Debridement Procedures | 1951 | 1953 |  |
|  | Anesthesia for Obstetric Procedures | 1958 | 1969 |  |
|  | Anesthesia for Other Procedures | 1990 | 1999 |  |
| Surgery | General Surgical Procedures | 10004 | 10021 |  |
|  | Surgical Procedures on the Integumentary System | 10030 | 19499 |  |
|  | Surgical Procedures on the Musculoskeletal System | 20100 | 29999 |  |
|  | Surgical Procedures on the Respiratory System | 30000 | 32999 |  |
|  | Surgical Procedures on the Cardiovascular System | 33016 | 37799 |  |
|  | Surgical Procedures on the Hemic and Lymphatic Systems | 38100 | 38999 |  |
|  | Surgical Procedures on the Mediastinum and Diaphragm | 39000 | 39599 |  |
|  | Surgical Procedures on the Digestive System | 40490 | 49999 |  |
|  | Surgical Procedures on the Urinary System | 50010 | 53899 |  |
|  | Surgical Procedures on the Male Genital System | 54000 | 55899 |  |
|  | Reproductive System Procedures | 55920 | 55920 |  |
|  | Intersex Surgery | 55970 | 55980 |  |
|  | Surgical Procedures on the Female Genital System | 56405 | 58999 |  |
|  | Surgical Procedures for Maternity Care and Delivery | 59000 | 59899 |  |
|  | Surgical Procedures on the Endocrine System | 60000 | 60699 |  |
|  | Surgical Procedures on the Nervous System | 61000 | 64999 |  |
|  | Surgical Procedures on the Eye and Ocular Adnexa | 65091 | 68899 |  |
|  | Surgical Procedures on the Auditory System | 69000 | 69979 |  |
|  | Operating Microscope Procedures | 69990 | 69990 |  |
| Radiology Procedures | Diagnostic Radiology (Diagnostic Imaging) Procedures | 70010 | 76499 |  |
|  | Diagnostic Ultrasound Procedures | 76506 | 76999 |  |
|  | Radiologic Guidance | 77001 | 77022 |  |
|  | Breast, Mammography | 77046 | 77067 |  |
|  | Bone/Joint Studies | 77071 | 77086 |  |
|  | Radiation Oncology Treatment | 77261 | 77799 |  |
|  | Nuclear Medicine Procedures | 78012 | 79999 |  |
| Pathology and Laboratory | Proprietary Laboratory Analyses | 1 | 247 | U |
|  | Organ or Disease Oriented Panels | 80047 | 80081 |  |
|  | Therapeutic Drug Assays | 80143 | 80377 |  |
|  | Drug Assay Procedures | 80305 | 83992 |  |
|  | Evocative/Suppression Testing Procedures | 80400 | 80439 |  |
|  | Clinical Pathology Consultations | 80500 | 80502 |  |
|  | Urinalysis Procedures | 81000 | 81099 |  |
|  | Molecular Pathology Procedures | 81105 | 81479 |  |
|  | Genomic Sequencing Procedures and Other Molecular Multianalyte Assays | 81410 | 81471 |  |
|  | Multianalyte Assays with Algorithmic Analyses | 81490 | 81599 |  |
|  | Chemistry Procedures | 82009 | 84999 |  |
|  | Hematology and Coagulation Procedures | 85002 | 85999 |  |
|  | Immunology Procedures | 86000 | 86849 |  |
|  | Transfusion Medicine Procedures | 86850 | 86999 |  |
|  | Microbiology Procedures | 87003 | 87999 |  |
|  | Anatomic Pathology Procedures | 88000 | 88099 |  |
|  | Cytopathology Procedures | 88104 | 88199 |  |
|  | Cytogenetic Studies | 88230 | 88299 |  |
|  | Surgical Pathology Procedures | 88300 | 88399 |  |
|  | In Vivo (eg, Transcutaneous) Laboratory Procedures | 88720 | 88749 |  |
|  | Other Pathology and Laboratory Procedures | 89049 | 89240 |  |
|  | Reproductive Medicine Procedures | 89250 | 89398 |  |
| Medicine Services and Procedure | Immune Globulins, Serum or Recombinant Products | 90281 | 90399 |  |
|  | Immunization Administration for Vaccines/Toxoids | 90460 | 90475 |  |
|  | Vaccines, Toxoids | 90476 | 90756 |  |
|  | Psychiatry Services and Procedures | 90785 | 90899 |  |
|  | Biofeedback Services and Procedures | 90901 | 90913 |  |
|  | Dialysis Services and Procedures | 90935 | 90999 |  |
|  | Gastroenterology Procedures | 91010 | 91299 |  |
|  | COVID-19 Vaccines/Toxoids | 91300 | 91304 |  |
|  | Ophthalmology Services and Procedures | 92002 | 92499 |  |
|  | Special Otorhinolaryngologic Services and Procedures | 92502 | 92700 |  |
|  | Cardiovascular Procedures | 92920 | 93799 |  |
|  | Non-Invasive Vascular Diagnostic Studies | 93880 | 93998 |  |
|  | Pulmonary Procedures | 94002 | 94799 |  |
|  | Allergy and Clinical Immunology Procedures | 95004 | 95199 |  |
|  | Endocrinology Services | 95249 | 95251 |  |
|  | Neurology and Neuromuscular Procedures | 95700 | 96020 |  |
|  | Medical Genetics and Genetic Counseling Services | 96040 | 96040 |  |
|  | Central Nervous System Assessments/Tests (eg, Neuro-Cognitive, Mental Status, Speech Testing) | 96105 | 96146 |  |
|  | Health Behavior Assessment and Intervention Procedures | 96156 | 96171 |  |
|  | Hydration, Therapeutic, Prophylactic, Diagnostic Injections and Infusions, and Chemotherapy and Other Highly Complex Drug or Highly Complex Biologic Agent Administration | 96360 | 96549 |  |
|  | Photodynamic Therapy Procedures | 96567 | 96574 |  |
|  | Special Dermatological Procedures | 96900 | 96999 |  |
|  | Physical Medicine and Rehabilitation Evaluations | 97010 | 97799 |  |
|  | Adaptive Behavior Services | 97151 | 97158 |  |
|  | Medical Nutrition Therapy Procedures | 97802 | 97804 |  |
|  | Acupuncture Procedures | 97810 | 97814 |  |
|  | Osteopathic Manipulative Treatment Procedures | 98925 | 98929 |  |
|  | Chiropractic Manipulative Treatment Procedures | 98940 | 98943 |  |
|  | Education and Training for Patient Self-Management | 98960 | 98962 |  |
|  | Non-Face-to-Face Nonphysician Services | 98966 | 98972 |  |
|  | Special Services, Procedures and Reports | 99000 | 99091 |  |
|  | Qualifying Circumstances for Anesthesia | 99100 | 99140 |  |
|  | Moderate (Conscious) Sedation | 99151 | 99157 |  |
|  | Other Medicine Services and Procedures | 99170 | 99199 |  |
|  | Home Health Procedures and Services | 99500 | 99602 |  |
|  | Medication Therapy Management Services | 99605 | 99607 |  |
| Evaluation and Management | Non-Face-to-Face Evaluation and Management Services | 99091 | 99474 |  |
|  | Office or Other Outpatient Services | 99202 | 99215 |  |
|  | Hospital Observation Services | 99217 | 99226 |  |
|  | Hospital Inpatient Services | 99221 | 99239 |  |
|  | Consultation Services | 99241 | 99255 |  |
|  | Emergency Department Services | 99281 | 99288 |  |
|  | Critical Care Services | 99291 | 99292 |  |
|  | Nursing Facility Services | 99304 | 99318 |  |
|  | Domiciliary, Rest Home (eg, Boarding Home), or Custodial Care Services | 99324 | 99337 |  |
|  | Domiciliary, Rest Home (eg, Assisted Living Facility), or Home Care Plan Oversight Services | 99339 | 99340 |  |
|  | Home Services | 99341 | 99350 |  |
|  | Prolonged Services | 99354 | 99417 |  |
|  | Case Management Services | 99366 | 99368 |  |
|  | Care Plan Oversight Services | 99374 | 99380 |  |
|  | Preventive Medicine Services | 99381 | 99429 |  |
|  | Care Management Evaluation and Management Services | 99439 | 99491 |  |
|  | Special Evaluation and Management Services | 99450 | 99458 |  |
|  | Newborn Care Services | 99460 | 99463 |  |
|  | Delivery/Birthing Room Attendance and Resuscitation Services | 99464 | 99465 |  |
|  | Inpatient Neonatal Intensive Care Services and Pediatric and Neonatal Critical Care Services | 99466 | 99486 |  |
|  | Cognitive Assessment and Care Plan Services | 99483 | 99486 |  |
|  | General Behavioral Health Integration Care Management | 99484 | 99484 |  |
|  | Psychiatric Collaborative Care Management Services | 99492 | 99494 |  |
|  | Transitional Care Evaluation and Management Services | 99495 | 99496 |  |
|  | Advance Care Planning Evaluation and Management Services | 99497 | 99498 |  |
|  | Other Evaluation and Management Services | 99499 | 99499 |  |
| Category II | Composite Measures | 1 | 15 | F |
|  | Patient Management | 500 | 584 | F |
|  | Patient History | 1000 | 1505 | F |
|  | Physical Examination | 2000 | 2060 | F |
|  | Diagnostic/Screening Processes or Results | 3006 | 3776 | F |
|  | Therapeutic, Preventive or Other Interventions | 4000 | 4563 | F |
|  | Follow-up or Other Outcomes | 5005 | 5250 | F |
|  | Patient Safety | 6005 | 6150 | F |
|  | Structural Measures | 7010 | 7025 | F |
|  | Non-Measure Category II Codes | 9001 | 9007 | F |
| Multianalyte | Multianalyte | 2 | 2 | M |
|  | Multianalyte | 3 | 3 | M |
|  | Multianalyte | 4 | 4 | M |
|  | Multianalyte | 6 | 6 | M |
|  | Multianalyte | 7 | 7 | M |
|  | Multianalyte | 11 | 11 | M |
|  | Multianalyte | 12 | 12 | M |
|  | Multianalyte | 13 | 13 | M |
|  | Multianalyte | 14 | 14 | M |
|  | Multianalyte | 15 | 15 | M |
|  | Multianalyte | 16 | 16 | M |
|  | Multianalyte | 17 | 17 | M |
| Category III | Various Services - Category III Codes | 42 | 184 | T |
|  | Remote Real-Time Interactive Video-conferenced Critical Care Services and Other Undefined Category Codes | 191 | 232 | T |
|  | Atherectomy (Open or Percutaneous) for Supra-Inguinal Arteries and Other Undefined Category Codes | 234 | 317 | T |
|  | Imaging, Testing, Implantation and Other Services | 329 | 358 | T |
|  | Adaptive Behavior Assessments | 362 | 373 | T |
|  | Other Procedures and Assessments | 376 | 379 | T |
|  | Pacemaker - Leadless and Pocketless System | 394 | 423 | T |
|  | Phrenic Nerve Stimulation System Procedures | 424 | 468 | T |
|  | Imaging, evaluation, programming and recording procedures | 469 | 478 | T |
|  | Laser ablation procedures | 479 | 480 | T |
|  | Blood products transfusion procedure | 481 | 481 | T |
|  | Cardiac diagnostic imaging and surgical procedures | 483 | 484 | T |
|  | Diagnostic procedures | 485 | 487 | T |
|  | Behavior Analysis | 488 | 488 | T |
|  | Cellular regeneration, evaluation study and ablation procedures | 489 | 493 | T |
|  | Organ transplantation procedures | 494 | 496 | T |
|  | Cardiac imaging procedures | 497 | 498 | T |
|  | Procedures performed on Urethra | 499 | 499 | T |
|  | Human Papillomavirus (HPV) analysis | 500 | 500 | T |
|  | Coronary artery disease (CAD) analysis | 501 | 504 | T |
|  | Other Diagnostic and Therapeutic Procedures | 505 | 508 | T |
|  | Vision Studies, Implants and Therapies | 509 | 514 | T |
|  | Cardiac Device Implantation, Analysis and Removal Procedures | 515 | 523 | T |
|  | Ablation Procedures | 524 | 524 | T |
|  | Intracardiac Ischemia Monitoring Procedures | 525 | 532 | T |
|  | Movement Disorder Analysis | 533 | 536 | T |
|  | Cellular Therapy Procedures | 537 | 540 | T |
|  | Cardiac Muscle Imaging | 541 | 542 | T |
|  | Cardiac Valve Repair Procedures | 543 | 545 | T |
|  | Radiofrequency Spectrometry Assessment and Bone Quality Testing Procedures | 546 | 547 | T |
|  | Incontinence Management Procedures | 548 | 551 | T |
|  | Laser Therapy and Implant Procedures | 552 | 553 | T |
|  | Bone Strength And Fracture Risk Assessment | 554 | 557 | T |
|  | Computed Tomography Analysis | 558 | 558 | T |
|  | Anatomic Model And Guide Creation | 559 | 562 | T |
|  | Chemo Drug Essay, Implant and Other Procedures | 563 | 568 | T |
|  | Cardiac Procedures with Evaluation on Valves and ICD System | 569 | 580 | T |
|  | Ablation Procedures | 581 | 582 | T |
|  | Procedures Peformed on Ear | 583 | 583 | T |
|  | Islet Cell Transplant Procedure | 584 | 586 | T |
|  | Neurostimulation Procedures | 587 | 590 | T |
|  | Health And Well-Being Coaching | 591 | 593 | T |
|  | Limb Lengthening Procedure | 594 | 594 | T |
|  | Female Voiding Prosthesis Procedures | 596 | 597 | T |
|  | Wound Imaging for Bacterial Presence | 598 | 599 | T |
|  | Irreversible Electroporation Ablation Procedures | 600 | 601 | T |
|  | Transdermal GFR Measurement and Monitoring | 602 | 603 | T |
|  | Eye Imaging Procedures (Remote) | 604 | 606 | T |
|  | Remote Monitoring of Pulmonary System | 607 | 608 | T |
|  | Magnetic Resonance Spectroscopy Imaging | 609 | 612 | T |
|  | Cardiac Implantation and Replacement Procedures | 613 | 614 | T |
|  | Procedures performed on Eyes | 615 | 618 | T |
|  | Procedures performed on Prostate | 619 | 619 | T |
|  | Endovascular Lower Limb Procedure | 620 | 620 | T |
|  | Trabeculostomy Procedure by Laser (Ab Interno) | 621 | 622 | T |
|  | Automated Analysis of Coronary Atherosclerotic plaque for CAD | 623 | 626 | T |
|  | Percutaneous Lumbar Intravertebral Disc Injection Procedure | 627 | 630 | T |
|  | Hyperspectral Imaging Measurement of Hemoglobin | 631 | 631 | T |
|  | Transcatheter Ultrasound Nerve Ablation Procedure | 632 | 632 | T |
|  | CT Breast (with/without Contrast) | 633 | 638 | T |
|  | CSF Shunt Analysis | 639 | 639 | T |

*Supplementary Table 1: CPT groups used as features; each group is defined by minimum and maximum numerical value of code and alphabetic modifier (if any).*

We used 21 most common CPT groups as features in our experiments. All of these groups had more than 1000 frequency in our dataset. Supplementary Table 2 shows selected CPT and their frequency values.

| **CPT Group Description** | **Count** |
| --- | --- |
| Hematology and Coagulation Procedures | 9111 |
| Organ or Disease Oriented Panels | 9099 |
| Drug Assay Procedures | 9014 |
| Chemistry Procedures | 8801 |
| Diagnostic Radiology (Diagnostic Imaging) Procedures | 8657 |
| Microbiology Procedures | 8603 |
| Cardiovascular Procedures | 8574 |
| Surgical Procedures on the Cardiovascular System | 7826 |
| Pulmonary Procedures | 7812 |
| Immunology Procedures | 7692 |
| Non-Face-to-Face Evaluation and Management Services | 6576 |
| Transfusion Medicine Procedures | 5284 |
| Hydration, Therapeutic, Prophylactic, Diagnostic Injections and Infusions, and Chemotherapy and Other Highly Complex Drug or Highly Complex Biologic Agent Administration | 4270 |
| Non-Invasive Vascular Diagnostic Studies | 4223 |
| Therapeutic Drug Assays | 3744 |
| Surgical Procedures on the Respiratory System | 3006 |
| Other Pathology and Laboratory Procedures | 2715 |
| Physical Medicine and Rehabilitation Evaluations | 2618 |
| Diagnostic Ultrasound Procedures | 2326 |
| Dialysis Services and Procedures | 2062 |
| Special Otorhinolaryngologic Services and Procedures | 1239 |

*Supplementary Table 2: Top-21 CPT groups in the dataset; each groups has more than 1000 frequency.*

#### ICD-9 Codes:

ICD-9 codes are very commonly used to represent comorbidities of patients in EMR. This ontology has hierarchical structure composed to groups and subgroups. We used each subgroup as a feature. Supplementary Table 3 shows the complete list of groups and subgroups in ICD-9 ontology. Similar feature engineering was applied in [CITE].

| **ICD-9 Start** | **ICD-9 End** | **Group Description** | **Subgroup Description** |
| --- | --- | --- | --- |
| 760 | 763.99 | Certain conditions originating in the perinatal period | Maternal causes of perinatal morbidity and mortality |
| 764 | 779.99 |  | Other conditions originating in the perinatal period |
| 640 | 640.99 | Complications of pregnancy, childbirth, and the puerperium | Complications mainly related to pregnancy |
| 660 | 669.99 |  | Complications occurring mainly in the course of labor and delivery |
| 670 | 677.99 |  | Complications of the puerperium |
| 630 | 633.99 |  | Ectopic and molar pregnancy |
| 650 | 659.99 |  | Normal delivery, and other indications for care in pregnancy, labor, and delivery |
| 678 | 679.99 |  | Other maternal and fetal complications |
| 634 | 639.99 |  | Other pregnancy with abortive outcome |
| 740 | 740 |  | Anencephalus and similar anomalies |
| 748 | 748 | Congenital anomalies | Anomalies of respiratory system, congenital |
| 745 | 745 |  | Bulbus cordis anomalies and anomalies of cardiac septal closure |
| 754 | 754 |  | Certain congenital musculoskeletal deformities |
| 758 | 758 |  | Chromosomal anomalies |
| 749 | 749 |  | Cleft palate and cleft lip |
| 744 | 744 |  | Congenital anomalies of ear, face, and neck |
| 743 | 743 |  | Congenital anomalies of eye |
| 752 | 752 |  | Congenital anomalies of genital organs |
| 757 | 757 |  | Congenital anomalies of the integument |
| 753 | 753 |  | Congenital anomalies of urinary system |
| 759 | 759 |  | Other and unspecified congenital anomalies |
| 747 | 747 |  | Other congenital anomalies of circulatory system |
| 751 | 751 |  | Other congenital anomalies of digestive system |
| 746 | 746 |  | Other congenital anomalies of heart |
| 755 | 755 |  | Other congenital anomalies of limbs |
| 742 | 742 |  | Other congenital anomalies of nervous system |
| 750 | 750 |  | Other congenital anomalies of upper alimentary tract |
| 756 | 756 |  | Other congenital musculoskeletal anomalies |
| 741 | 741 |  | Spina bifida |
| 283 | 283 |  | Acquired hemolytic anemias |
| 284 | 284 | Diseases of the blood and the blood forming organs | Aplastic anemia and other bone marrow failure syndromes |
| 286 | 286 |  | Coagulation defects |
| 288 | 288 |  | Diseases of white blood cells |
| 282 | 282 |  | Hereditary hemolytic anemias |
| 280 | 280 |  | Iron deficiency anemias |
| 285 | 285 |  | Other and unspecified anemias |
| 281 | 281 |  | Other deficiency anemias |
| 289 | 289 |  | Other diseases of blood and blood-forming organs |
| 287 | 287 |  | Purpura and other hemorrhagic conditions |
| 390 | 392.99 |  | Acute rheumatic fever |
| 430 | 438.99 | Diseases of the circulatory system | Cerebrovascular disease |
| 393 | 398.99 |  | Chronic rheumatic heart disease |
| 440 | 449.99 |  | Diseases of arteries, arterioles, and capillaries |
| 415 | 417.99 |  | Diseases of pulmonary circulation |
| 451 | 459.99 |  | Diseases of veins and lymphatics, and other diseases of circulatory system |
| 401 | 405.99 |  | Hypertensive disease |
| 410 | 414.99 |  | Ischemic heart disease |
| 420 | 429.99 |  | Other forms of heart disease |
| 540 | 543.99 | Diseases of the digestive system | Appendicitis |
| 530 | 539.99 |  | Diseases of esophagus, stomach, and duodenum |
| 520 | 529.99 |  | Diseases of oral cavity, salivary glands, and jaws |
| 538 | 538 |  | Gastrointestinal mucositis (ulcerative) |
| 550 | 553.99 |  | Hernia of abdominal cavity |
| 555 | 558.99 |  | Noninfectious enteritis and colitis |
| 570 | 579.99 |  | Other diseases of digestive system |
| 560 | 569.99 |  | Other diseases of intestines and peritoneum |
| 600 | 608.99 | Diseases of the genitourinary system | Diseases of male genital organs |
| 610 | 612.99 |  | Disorders of breast |
| 614 | 616.99 |  | Inflammatory disease of female pelvic organs |
| 580 | 589.99 |  | Nephritis, nephrotic syndrome, and nephrosis |
| 590 | 599.99 |  | Other diseases of urinary system |
| 617 | 629.99 |  | Other disorders of female genital tract |
| 710 | 719.99 | Diseases of the musculoskeletal system and connective tissue | Arthropathies and related disorders |
| 720 | 724.99 |  | Dorsopathies |
| 730 | 739.99 |  | Osteopathies, chondropathies, and acquired musculoskeletal deformities |
| 725 | 729.99 |  | Rheumatism, excluding the back |
| 380 | 389.99 | Diseases of the nervous system and sense organs | Diseases of the ear and mastoid process |
| 360 | 379.99 |  | Disorders of the eye and adnexa |
| 350 | 359.99 |  | Disorders of the peripheral nervous system |
| 330 | 337.99 |  | Hereditary and degenerative diseases of the central nervous system |
| 320 | 326.99 |  | Inflammatory diseases of the central nervous system |
| 327 | 327.99 |  | Organic sleep disorders |
| 340 | 349.99 |  | Other disorders of the central nervous system |
| 339 | 339.99 |  | Other headache syndromes |
| 338 | 338.99 |  | Pain |
| 460 | 466.99 |  | Acute respiratory infections |
| 490 | 496.99 | Diseases of the respiratory system | Chronic obstructive pulmonary disease and allied conditions |
| 510 | 519.99 |  | Other diseases of respiratory system |
| 470 | 478.99 |  | Other diseases of the upper respiratory tract |
| 500 | 508.99 |  | Pneumoconiosis and other |
| 480 | 488.99 |  | Lung diseases due to external agents |
| 680 | 686.99 |  | Pneumonia and influenza |
| 700 | 709.99 | Diseases of the skin and subcutaneous tissue | Other diseases of skin and subcutaneous tissue |
| 690 | 698.99 |  | Other inflammatory conditions of skin and subcutaneous tissue |
| 249 | 259.99 | Endocrine, nutritional and metabolic diseases, and immunity disorders | Diseases of other endocrine glands |
| 240 | 246.99 |  | Disorders of thyroid gland |
| 260 | 269.99 |  | Nutritional deficiencies |
| 270 | 279.99 |  | Other metabolic and immunity disorders |
| 60 | 66.99 | Infectious and parasitic diseases | Arthropod-borne viral diseases |
| 120 | 129.99 |  | Helminthiases |
| 42 | 42.99 |  | Human immunodeficiency |
| 1 | 9.99 |  | Virus [HIV] infection |
| 137 | 139.99 |  | Intestinal infectious diseases |
| 110 | 118.99 |  | Late effects of infectious and parasitic diseases |
| 30 | 41.99 |  | Mycoses |
| 70 | 79.99 |  | Other bacterial diseases |
| 130 | 136.99 |  | Other diseases due to viruses and chlamydia |
| 100 | 104.99 |  | Other infectious and parasitic diseases |
| 45 | 49.99 |  | Other spirochetal diseases |
| 80 | 88.99 |  | Poliomyelitis and other non-arthropod-borne viral diseases and prion diseases of central nervous system |
| 90 | 99.99 |  | Rickettsioses and other arthropod-borne diseases |
| 10 | 18.99 |  | Syphilis and other venereal diseases |
| 50 | 59.99 |  | Tuberculosis |
| 20 | 27.99 |  | Viral diseases generally accompanied by exanthem |
| 940 | 949.99 | Injury and poisoning | Burns |
| 958 | 959.99 |  | Certain traumatic complications and unspecified injuries |
| 996 | 999.99 |  | Complications of surgical and medical care, not elsewhere classified |
| 920 | 924.99 |  | Contusion with intact skin surface |
| 925 | 929.99 |  | Crushing injury |
| 830 | 839.99 |  | Dislocation |
| 930 | 939.99 |  | Effects of foreign body |
| 800 | 829.99 |  | Entering through orifice fractures |
| 900 | 904.99 |  | Injury to blood vessels |
| 950 | 957.99 |  | Injury to nerves and spinal cord |
| 860 | 869.99 |  | Internal injury of thorax, abdomen, and pelvis |
| 850 | 854.99 |  | Intracranial injury, excluding those with skull fracture |
| 905 | 909.99 |  | Late effects of injuries, poisonings, toxic effects, and other external causes |
| 870 | 897.99 |  | Open wounds |
| 990 | 995.99 |  | Other and unspecified effects of external causes |
| 960 | 979.99 |  | Poisoning by drugs, medicinal and biological substances |
| 840 | 848.99 |  | Sprains and strains of joints and adjacent muscles |
| 910 | 919.99 |  | Superficial injury |
| 980 | 989.99 |  | Toxic effects of substances chiefly nonmedicinal as to source |
| 317 | 319.99 |  | Intellectual disabilities |
| 300 | 316.99 | Mental, behavioral and neurodevelopmental disorders | Neurotic disorders, personality disorders, and other nonpsychotic mental disorders |
| 290 | 299.99 |  | Psychoses |
| 140 | 239.99 | Neoplasms | Neoplasms |
| 797 | 799.99 |  | Ill-defined and unknown causes of morbidity and mortality |
| 790 | 796.99 | Symptoms, signs, and ill-defined conditions | Nonspecific abnormal findings |
| 780 | 780 |  | General symptoms |
| 789 | 789 |  | Other symptoms involving abdomen and pelvis |
| 783 | 783 |  | Symptoms concerning nutrition, metabolism, and development |
| 785 | 785 |  | Symptoms involving cardiovascular system |
| 787 | 787 |  | Symptoms involving digestive system |
| 784 | 784 |  | Symptoms involving head and neck |
| 781 | 781 |  | Symptoms involving nervous and musculoskeletal systems |
| 786 | 786 |  | Symptoms involving respiratory system and other chest symptoms |
| 782 | 782 |  | Symptoms involving skin and other integumentary tissue |
| 788 | 788 | Symptoms, signs, and ill-defined conditions | Symptoms involving urinary system |
| E000 | E999.99 |  | Supplementary classification of external causes of injury and poisoning |
| V01 | V91.99 | Supplementary classification of factors influencing health status and contact with health services | Supplementary classification of factors influencing health status and contact with health services |

*Supplementary Table 3: ICD-9 ontology; each subgroup is used as feature.*

We selected top 30 subgroups, based on frequency in our dataset, in our experiment. Supplementary Table 4 shows these subgroups and their frequencies.

| **Count** | **ICD Subgroup Name** |
| --- | --- |
| 122997 | SUPPLEMENTARY CLASSIFICATION OF FACTORS INFLUENCING HEALTH STATUS AND CONTACT WITH HEALTH SERVICES |
| 34963 | OTHER METABOLIC AND IMMUNITY DISORDERS |
| 34841 | OTHER DISEASES OF RESPIRATORY SYSTEM |
| 32802 | OTHER FORMS OF HEART DISEASE |
| 31993 | NEPHRITIS, NEPHROTIC SYNDROME, AND NEPHROSIS |
| 28978 | HYPERTENSIVE DISEASE |
| 22975 | DISEASES OF OTHER ENDOCRINE GLANDS |
| 20976 | NONSPECIFIC ABNORMAL FINDINGS |
| 18555 | NEOPLASMS |
| 16916 | Symptoms involving respiratory system and other chest symptoms |
| 13782 | General symptoms |
| 13031 | LUNG DISEASES DUE TO EXTERNAL AGENTS |
| 9101 | NEUROTIC DISORDERS, PERSONALITY DISORDERS, AND OTHER NONPSYCHOTIC MENTAL DISORDERS |
| 8931 | Other and unspecified anemias |
| 8192 | Symptoms involving digestive system |
| 8026 | ARTHROPATHIES AND RELATED DISORDERS |
| 7964 | OTHER DISORDERS OF THE CENTRAL NERVOUS SYSTEM |
| 7664 | ISCHEMIC HEART DISEASE |
| 7151 | OTHER DISEASES OF URINARY SYSTEM |
| 7113 | Symptoms involving cardiovascular system |
| 7111 | CHRONIC OBSTRUCTIVE PULMONARY DISEASE AND ALLIED CONDITIONS |
| 6576 | DISEASES OF VEINS AND LYMPHATICS, AND OTHER DISEASES OF CIRCULATORY SYSTEM |
| 6429 | MYCOSES |
| 6422 | OTHER DISEASES OF DIGESTIVE SYSTEM |
| 5893 | DISEASES OF ESOPHAGUS, STOMACH, AND DUODENUM |
| 5306 | Other symptoms involving abdomen and pelvis |
| 5227 | DORSOPATHIES |
| 4997 | CEREBROVASCULAR DISEASE |
| 4864 | PSYCHOSES |

*Supplementary Table 4: ICD-9 subgroups included in our experiments*

### Model Design:

#### Fusion Model:

Graph convolutional neural networks (GCN) present an intuitive and elegant way of processing multi-modal data presented as a graph structure. GCN learns to generate vector embedding of each node optimized for the downstream prediction task, like any convolutional neural network (CNN). While CNN defines ‘neighborhood’ of an instance based on spatial proximity such as neighboring pixel in images, GCN allows neighborhood to be defined by edges in graph structure. Thus node embeddings generated by a GCN are dependent not only on node representation but also representation of neighboring/edge-connected nodes. This arrangement results in better node embeddings if definition of neighborhood, i.e., edges, is meaningful. As described earlier, we form graph structure by using chest X-rays as nodes, and define edges based on similarity in patients’ demographic information or medical history. Therefore, the notion of ‘neighborhood’ is based on useful clinical information while making predictions for future clinical events.

#### Time Threshold for Prediction:

The goal of our project is advanced prediction of clinical events to facilitate hospital resource management. Generally such prediction is limited to 24 hours, 48 hours or 72 hours in advance. We investigated our training cohort and experiments with 24 hours upto 7 days time thresholds. Our experiments led us to 72 hours or 3 days as the time threshold. Prediction label distribution is highly imbalanced, but the 3 days threshold provides the most balanced distribution of discharged ns. not discharged labels as shown in Supplementary Figure 1. For the sake of consistency in the overall pipeline, we used the same threshold for the remaining two decision time points.

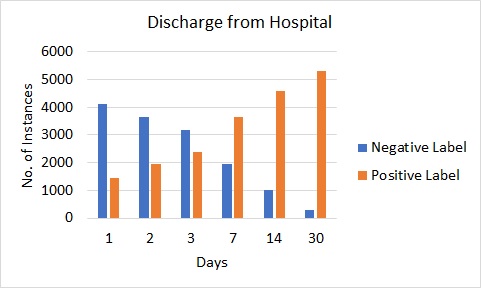

*Supplementary Figure 1: Distribution of discharged and not-discharged labels with different prediction time thresholds*

#### Branched Framework vs. Multiclass Predictors

Our current framework is based on a branched pipeline, consisting of three binary predictors. While designing this framework, we experimented with both multiple binary predictors as well as multi-class predictors. The challenge for multi-class classifiers is the extreme imbalance of class distribution. Some class labels such as ‘admission to ICU’ and ‘death’ of the patient have much smaller representations than classes like ‘discharged’ from hospital and ‘Not discharged’ from hospital. A small fraction of patients from ‘Not discharged’ from hospital are admitted to ICU. Similarly, a small fraction of patients who are ‘discharged’ from hospital, expire within the defined time-period.

In this section, we present the results of two multi-class classifiers; a) Classifier01- prediction labels ‘discharged’, ‘not discharged’, ‘expired’ with the given time period of three days, and b) Classifier02- prediction labels ‘discharged’, ‘not discharged’, ‘admitted to ICU’ within the given time periods of three days. Note the ‘discharged’ label for Classifier01 is used for samples who are discharged from hospital, but do not expire. Similarly, the ‘not discharged’ label for Classifier02 is used for samples who remain in hospital, but are not admitted to ICU. We included random subsampling to counter the class imbalance. The best results are shown in Supplementary Tables 5 and 6.

*Supplementary Table 5: Performance of classifier01 for graphs based on different information sources from EMR*

|  |  | **Precision** | **Recall** | **F1-score** | **Number of samples** |
| --- | --- | --- | --- | --- | --- |
| **Demographics-based graph** | Discharged | 70 | 60 | 65 | 445 |
|  | Not Discharged | 75 | 89 | 82 | 982 |
|  | Dead | 0 | 0 | 0 | 118 |
|  | Overall | 48 | 50 | 49 |  |
| **CPT-graph** | Discharged | 62 | 64 | 63 | 445 |
|  | Not Discharged | 76 | 83 | 79 | 982 |
|  | Dead | 0 | 0 | 0 | 118 |
|  | Overall | 46 | 49 | 47 |  |
| **ICD 9-based graph** | Discharged | 56 | 64 | 59 | 445 |
|  | Not Discharged | 80 | 68 | 73 | 982 |
|  | Dead | **10** | **16** | **12** | 118 |
|  | Overall | 48 | 49 | 48 |  |

*Supplementary Table 6: Performance of classifier02 for graphs based on different information sources from EMR*

|  |  | **Precision** | **Recall** | **F1-score** | **Number of samples** |
| --- | --- | --- | --- | --- | --- |
| **Demographics-based graph** | Discharged | 79 | 40 | 53 | 563 |
|  | Not Discharged | 73 | 79 | 76 | 872 |
|  | Admitted to ICU | 12 | 34 | 18 | 110 |
|  | Overall | 54 | 51 | 49 |  |
| **CPT-graph** | Discharged | 77 | 40 | 53 | 563 |
|  | Not Discharged | 71 | 88 | 78 | 872 |
|  | Admitted to ICU | **18** | **29** | **22** | 110 |
|  | Overall | 55 | 52 | 51 |  |
| **ICD 9-based graph** | Discharged | 62 | 42 | 50 | 563 |
|  | Not Discharged | 71 | 80 | 75 | 872 |
|  | Admitted to ICU | 15 | 25 | 19 | 110 |
|  | Overall | 49 | 49 | 48 |  |

Note that the performance of minority class, i.e., ‘Dead’ and ‘Admitted to ICU’ is very poor. With our multiple binary predictors framework, we were able to achieve 67% F1-score for ‘Dead’ label (compared to 12% F- score achieved by Classifier01). Similarly, the binary predictor was able to achieve a 46% F1-score for ‘Admitted to ICU’ (compared with 22% F1-score achieved by Classifier02).

With such disparity in performance, we decided to use a pipeline consisting of multiple binary predictors.

#### Threshold Selection for Edge Formation:

Edges between chest X-rays are based on demographics and medical history of the corresponding patient collected upto the time of X-ray study. Most of these features are categorical (continuous value feature ‘Age’ is binned to be converted to categorical values). We used thresholding on cosine similarity between feature vectors to decide edge formation. We noticed that decreasing the threshold results in a larger number of edges, increased processing time, but no significant performance improvement. The following Tables (supplementary Table 7, 8, and 9) show results of the best performing model configuration for each decision time-point models for three different threshold values. Threshold value of 0.9 was generally the best performing value.

*Supplementary Table 7: Performance of Model-1 with CPT based graph for different threshold values for edge formation*

| **Threshold** | **No. of edges** | **Avg. degree** |  | **Precision** | **Recall** | **F1-score** | **Number of samples** |
| --- | --- | --- | --- | --- | --- | --- | --- |
| **0.75** | 21350872 | 2732 | Discharged | 65 | 52 | 58 | 563 |
|  |  |  | Not Discharged | 75 | 84 | 80 | 982 |
|  |  |  | Overall | 70 | 68 | 69 |  |
| **0.9** | 3417408 | 437 | Discharged | 74 | 54 | 62 | 563 |
|  |  |  | Not Discharged | 77 | 89 | 83 | 982 |
|  |  |  | Overall | **76** | **72** | **73** |  |
| **1.0** | 124556 | 16 | Discharged | 62 | 60 | 61 | 563 |
|  |  |  | Not Discharged | 77 | 79 | 78 | 982 |
|  |  |  | Overall | 70 | 69 | 70 |  |

*Supplementary Table 8: Performance of Model-2 with CPT based graph for different threshold values for edge formation*

| **Threshold** | **No. of edges** | **Avg. degree** |  | **Precision** | **Recall** | **F1-score** | **Number of samples** |
| --- | --- | --- | --- | --- | --- | --- | --- |
| **0.75** | 1021488 | 345 | Alive | 93 | 85 | 89 | 445 |
|  |  |  | Dead | 57 | 75 | 65 | 118 |
|  |  |  | Overall | 75 | 80 | 77 |  |
| **0.9** | 142558 | 48 | Alive | 91 | 92 | 91 | 445 |
|  |  |  | Dead | 68 | 65 | 67 | 118 |
|  |  |  | Overall | **80** | **79** | **79** |  |
| **1.0** | 8780 | 3 | Alive | 89 | 91 | 90 | 445 |
|  |  |  | Dead | 62 | 58 | 60 | 118 |
|  |  |  | Overall | 75 | 74 | 75 |  |

*Supplementary Table 9: Performance of Model-3 with ICD-9 based graph for different threshold values for edge formation*

| **Threshold** | **No. of edges** | **Avg. degree** |  | **Precision** | **Recall** | **F1-score** | **Number of samples** |
| --- | --- | --- | --- | --- | --- | --- | --- |
| **0.75** | 912360 | 305 | Not Admitted to ICU | 88 | 74 | 81 | 482 |
|  |  |  | Admitted to ICU | 34 | 56 | 42 | 110 |
|  |  |  | Overall | 61 | 65 | 61 |  |
| **0.9** | 38442 | 13 | Not Admitted to ICU | 88 | 85 | 86 | 482 |
|  |  |  | Admitted to ICU | 43 | 50 | 46 | 110 |
|  |  |  | Overall | **65** | **67** | **66** |  |
| **1.0** | 13196 | 4 | Not Admitted to ICU | 86 | 87 | 86 | 482 |
|  |  |  | Admitted to ICU | 39 | 35 | 37 | 110 |
|  |  |  | Overall | 62 | 61 | 52 |  |

#### Transference Performance:

Model 1 at decision time point 1 is trained in a comprehensive set containing all instances in the data while the remaining two models are trained on subsets of the dataset. Model-1 is trained to predict a broad category of hospital discharge. The remaining two models predict labels as subcategories (ICU admission for patients not being discharged from the hospital, and mortality among patients discharged from hospital). We experimented with transferring Model-1 to the remaining two decision time points for prediction, and finetuning it further for the specific labels at each of those decision timepoints. Supplementary Table 10 shows performance of such model transference.

*Supplementary Table 10: Transfer performance of model*

| **Training decision-point** | **Testing decision-point** | Tuning |  | **Precision** | **Recall** | **F1-score** | **Number of samples** |
| --- | --- | --- | --- | --- | --- | --- | --- |
| **Decision point#1** | Decision point#2 | No | Alive | 90 | 92 | 91 | 445 |
|  |  |  | Dead | 67 | 62 | 64 | 118 |
|  |  |  | Overall | 79 | 77 | 78 |  |
| **Decision point#1** | Decision point#2 | Yes | Alive | 93 | 90 | 92 | 445 |
|  |  |  | Dead | 67 | 75 | 70 | 118 |
|  |  |  | Overall | 80 | 82 | 81 |  |
| **Decision point#1** | Decision point#3 | No | Not Admitted to ICU | 73 | 51 | 60 | 482 |
|  |  |  | Admitted to ICU | 8 | 18 | 11 | 110 |
|  |  |  | Overall | 41 | 35 | 36 | 592 |
| **Decision point#1** | Decision point#3 | Yes | Not Admitted to ICU | 86 | 83 | 84 | 482 |
|  |  |  | Admitted to ICU | 35 | 40 | 37 | 110 |
|  |  |  | Overall | 60 | 61 | 61 | 592 |

There are interesting implications for such transference approach. Even without finetuning, Model-1 is almost as good as a model trained solely on data at decision timepoint 2 for mortality prediction. After finetuning, such transferred model is able to outperform on a locally trained model. However, ICU admission is better predicted by using locally trained model, even though transferred model comes close to the performance of locally trained model after finetuning.

### External Validation

We selected a small cohort of COVID-19 patients from Mayo Clinic to perform external validation of our fusion based models. The performance of this validation is reported in the main manuscript. We would like to point out that distribution of medical codes such as CPT and ICD-10 are very different between our internal and external test sets. Supplementary Table 2 and 4 show common CPT and ICD-9 subgroups for internal data. Supplementary Tables 11 and 12 show common CPT and ICD-10 (comorbidities in the external set have been recorded as ICD-10 and not ICD-9 as was done in our internal test set) subgroups in the external test set. It is quite clear that common procedures and common comorbidities for the two sets are very different. However, one advantage of graph neural networks is that it requires only a network/graph of instances to operate on, leaving the formation of the graph as a design choice at the preprocessing stage with no impact on the applicability of the pretrained GNN. When we formed graphs of instances in the external set using common procedures and comorbidities of the external set, pretrained GNN was able to achieve decent performance. The baseline EHR-only model requires exactly the same comorbidities and procedures to be used as features for an external set for the model to be applicable on the external data.

| **Description of CPT Subgroups** | **Count** |
| --- | --- |
| Diagnostic Radiology (Diagnostic Imaging) Procedures | 241 |
| Surgical Procedures on the Cardiovascular System | 235 |
| Hematology and Coagulation Procedures | 232 |
| Organ or Disease Oriented Panels | 230 |
| Non-Face-to-Face Evaluation and Management Services | 228 |
| Drug Assay Procedures | 212 |
| Chemistry Procedures | 197 |
| Cardiovascular Procedures | 195 |
| Immunology Procedures | 194 |
| Microbiology Procedures | 189 |
| Pulmonary Procedures | 173 |
| Physical Medicine and Rehabilitation Evaluations | 134 |
| Diagnostic Ultrasound Procedures | 127 |
| Transfusion Medicine Procedures | 117 |
| Non-Invasive Vascular Diagnostic Studies | 107 |
| Surgical Procedures on the Respiratory System | 103 |

*Supplementary Table 11: Top-16 CPT groups in the dataset; each group has more than 100 frequencies.*

| **Description of ICD-10 Subgroup** | **Count** |
| --- | --- |
| Abnormal findings on diagnostic imaging and in function studies, without diagnosis | 204 |
| Other diseases of the respiratory system | 203 |
| Persons with potential health hazards related to family and personal history and certain conditions influencing health status | 201 |
| Symptoms and signs involving the circulatory and respiratory systems | 182 |
| Influenza and pneumonia | 172 |
| Provisional assignment of new diseases of uncertain etiology or emergency use | 167 |
| Encounters for other specific health care | 140 |
| Other respiratory diseases principally affecting the interstitium | 136 |
| Other forms of heart disease | 125 |
| Other disorders of the nervous system | 120 |
| Abnormal findings on examination of blood, without diagnosis | 119 |
| Metabolic disorders | 115 |
| General symptoms and signs | 106 |

*Supplementary Table 12: Top-13 CPT groups in the dataset; each group has more than 100 frequencies.*
